# A steady trickle-down from metro districts and improving epidemic-parameters characterized the increasing COVID-19 cases in India

**DOI:** 10.1101/2020.09.28.20202978

**Authors:** Santosh Ansumali, Aloke Kumar, Samarth Agarwal, H. J. Shashank, Meher K. Prakash

## Abstract

**Background:** By mid-September of 2020, the number of daily new infections in India crossed 95, 000. We aimed to characterize the spatio-temporal shifts in the disease burden as the infections rose during the first wave of COVID-19.

**Methods:** We gathered the publicly available district-level (equivalent of counties) granular data for the 15 April to 31 August 2020 period. We used the epidemiological data from 186 districts with the highest case burden as of August 31, 559, 566 active cases and 2, 715, 656 cumulative infections, and the governing epidemic parameters were estimated by fitting it to a susceptible-asymptomatic-infected-recovered-dead (SAIRD) model. The space-time trends in the case burden and epidemic parameters were analyzed. When the physical proximity of the districts did not explain the spreading patterns, we developed a metric for accessibility of the districts via air and train travel. The districts were categorized as large metro, metro, urban and sub-urban and the spatial shifts in case burden were analyzed.

**Results:** The center of the burden of the current-active infections which on May 15 was in the large metro districts with easy international access shifted continuously and smoothly towards districts which could be accessed by domestic airports and by trains. A linear trend-analysis showed a continuous improvement in the governing epidemic parameters consistently across the four categories of districts. The reproduction numbers improved from 1.77 ± 0.58 on May 15 to 1.07 ± 0.13 on August 31 in large metro districts (p-Value of trend 0.0001053); and from 1.58 ± 0.39 on May 15 to 0.94 ± 0.11 on August 31 in sub-urban districts (p-Value of trend 0.0067). The recovery rate per infected person per day improved from 0.0581 ± 0.009 on May 15 to 0.091 ± 0.010 on August 31 in large metro districts (p-Value of trend 0.26 × 10^−12^); and from 0.059 ± 0.011 on May 15 to 0.100 ± 0.010 on August 31 in sub-urban districts (p-Value of trend 0.12 × 10^−16^). The death rate of symptomatic individuals which includes the case-fatality-rate as well as the time from symptoms to death, consistently decreased from 0.0025 ± 0.0014 on May 15 to 0.0013 ± 0.0003 on August 31 in large metro districts (p-Value of trend 0.0010); and from 0.0018 ± 0.0008 on May 15 to 0.0014 ± 0.0003 on August 31 in sub-urban districts (p-Value of trend 0.2789).

**Conclusions:** As the daily infections continued to rise at a national level, the “center” of the pandemic-burden shifted smoothly and predictably towards smaller sized districts in a clear hierarchical fashion of accessibility from an international travel perspective. This observed trend was meant to serve as an alert to re-organize healthcare resources towards remote districts. The geographical spreading patterns continue to be relevant as the second wave of infections began in March 2021 with a center in the mid-range districts.

**Funding:** None

## Introduction

In the COVID-19 pandemic that began in December 2019, each month witnessed the critical rise of infections in completely different countries. The initial declaration of national emergency which was a “one-size fits all” was quickly irrelevant in many countries as each state or county perceived the need for stringency differently. The differences arose mainly because even within the first wave of the pandemic, the solidarity extended through interstate loans of ventilators demonstrated how at any given time the geographical regions in different phases of the pandemic, rising, stable or declining, perceived the threat differently from others. As the rise of infections in different geographical regions is asynchronous, the critical care burden shifts dynamically with each region attempting to achieve its ‘local-flattening’ of the peak at a different time. The focus shifted towards regional containment COVID-19 containment strategies, for which identifying the next hotspots became extremely important[1, 2].

Epidemiological models have focused on making predictions of the rise of infections, and a peak of critical care burden on a country or a state basis. There have been longitudinal studies[3, 4] demonstrating how the reproduction number varied over time in the different states or countries and possibly correlating it to the effects of lockdowns. Further, a few cross-sectional studies correlated the hotspots in the spread of infections in the initial phase to the traffic flow out of Wuhan.[5] These studies mostly focused on one region at a time, either on estimating parameters, or making projections. In this work, we build a summarizing perspective of the geographical development of the pandemic, by analyzing a cumulative 2.71 million infections and 52 thousand deaths of COVID-19 from India over a period of four and a half months.

To examine how the hotspots of COVID-19 spread over time across India, we identified the districts of interest based on case-burden, estimated the time-varying epidemiological parameters for these regions, categorized these regions according to the ease of travel access in the hierarchical order of availability of international flights, domestic flights, trains or other quaternary modes of transport, and analyzed the trends in parameters over time in each of these categories. The data shows predictable shifts to remote regions. The infections spreading to newer and under-catered locations presents both an opportunity and a challenge, an asynchronous peak of critical requirements which can be addressed both by manufacturing and re-organizing the resources on an *ad hoc* basis.

### Research in context

#### Evidence before this study

We searched *pubmed* and *medRxiv* up to 20 September 2020 with the keywords “COVID-19”, “time variation or longitudinal”, “cantons or states”, “hotspots”, “diffusion” “active-cases”. The published research thus far focused mainly on reporting either the time variation of reproduction number at the state-level, or the predictions of rise of infections at the canton level. However, from these constantly updating predictions with new hotspots appearing, there is no continuity of knowledge and possibility to obtain a comprehensive picture of the development of the pandemic. A study correlating the number of infections in different regions in China to the traffic-flow out of Wuhan, and another describing the development of hotspots along the highways in Brazil describe some aspects of this geographical spread. Firstly, a similar analysis with granular Indian data has been missing. Further, the questions of how the center of the active-case burden diffused and the spatio-temporal trends in epidemiological parameters underlying the large number of infections have not been the focus of these earlier studies.

#### Added value of this study

Our study begins with the understanding that the different geographic regions such as districts (or counties) may be in different phases of the pandemic, and the mixing of population within a district is more likely than across the different districts. Considering this geographical heterogeneity, and the ease of access to these different districts, we develop a summary of how the pandemic evolved in the regions of different accessibility over a period of four-and-a-half months. Predictable trends in the case-burden diffusion are identified.

#### Implications of all the available evidence

Authorities should understand the shift in the dynamics of the critical-care requirements, not by geographically contiguous regions, but by ease of travel or by common economic interests which guided such ease of travel. Strategies should be driven by knowing where the pandemic burden is likely to move, before it happens. The trend of the case-burden predictably shifting towards suburban districts can be useful in re-organizing the available resources as per the needs.

## Methods

### COVID-19 Data

The district-wise COVID-19 daily new infections, daily active cases and daily deaths were gathered from a publicly available data repository: *http://www.covid19india.org.* The age, sex demographics of the infected people in these districts were not available. Using a criterion of at least 20 daily new infections on 31 August 2020, 186 districts were identified and used in the analysis. These 186 districts accounted for 2, 715, 656 of the 3, 687, 939 cumulative infections in India, and had an active case burden of 559, 566 from the 785, 127 total active infections in India. These 186 districts out of the 739 districts in India, account for a 696 million population in 2020, estimated from the 2001 and 2011 census data. The data spans across 22 different states which are currently governed by many different political parties, which is likely to reduce any inherent and centralized biases in the data.

### Model

An important feature of COVID-19 is the role played by asymptomatics in the spread of the disease.[6] Classic compartmental models such as the susceptible infected-recovered (SIR) models fail to account for this important aspect of the disease and thus one should work with models where asymptomatic fraction is explicitly included in the model. In this work, we extended the susceptible-asymptomatic-infected-recovered (SAIR) model we developed earlier[7] by including the deaths (D), and used this SAIRD model to estimate the underlying epidemic parameters for each district. The compartmental nature of the model is shown schematically in Figure (1A) and the governing equations are given in Figure (1B). The asymptomatics in this framework are assumed to include the pre-symptomatics who subsequently become symptomatic as well as those who will continue to remain asymptomatic until their recovery. The rates of recovery and transmission of infections of the asymptomatic and the symptomatic individuals are assumed to be the same. The model thus relies on four different epidemiological parameters - infection rate (*β*), recovery rate (*γ*), death rate (*γ*_*D*_), asymptomatic to symptomatic conversion (*δ*), which we extracted from the data as follows.

**Figure 1:**
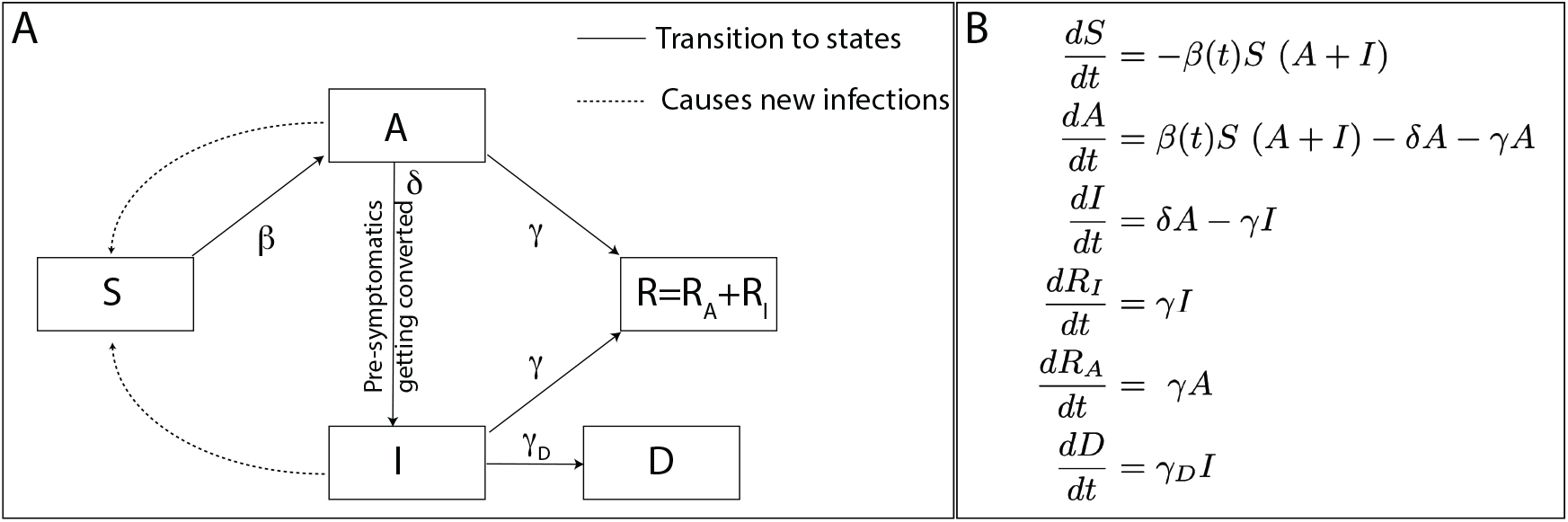
*A*. A schematic of the SAIRD model and its *B*. modelling equations used in the present analysis. The SAIR model on which this scheme is based is described in Ref. [7]. The parameters used in these equations are assumed to be constant over different segments of the pandemic trajectory (piece-wise continuous) and were estimated by fitting the predictions for infections, recoveries, and deaths over time.

### Parameter Estimation

Even in the simplest scenario of the exponential growth with of infections *I* with time, we found that choosing a linear fit between *dI/dt* and *I* led to a better parameter estimation than an exponential fit between *I* and *t*, although both are mathematically equivalent.[8] The parameter estimation scheme we evolved[7] for all the parameters thus relies on rearranging the underlying equations in a way that allowed us to provide most reliable estimates despite the uncertainties and noise in the data. The time-varying epidemiological parameters were estimated for the different districts. The infection spread-rate (*β*), recovery-rate (*γ*) and the death-rate (*γ*_*D*_) were estimated in a time-dependent way, identifying time periods over which a single parameter best describes the observations. The (pre-or) asymptomatic to symptomatic conversion rate (*δ*) was assumed to be a constant for each district.

*γ*(*t*) **estimation**. To estimate *γ* we note that, *dR*_*I*_*/dt* = *γI*, which can be easily integrated to obtain 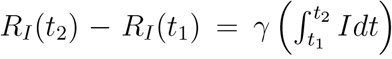. However, it is well understood that *γ* changes over time owing to changes in policies and behavioral patterns. We note that the plot for *R*_*I*_ vs. *Idt* is piecewise linear and the value of *γ* for different periods can be calculated using simple linear regression and estimating the slope.

*γ*_*D*_(*t*) **estimation**. The estimation for *γ*_*D*_ then follows in a similar procedure. *dD dt* = *γ_D_I*, which is integrated to obtain 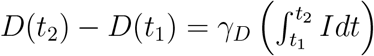.

*β*(*t*) **estimation**. To estimate *β* we employ log *I*(*t*) *∼* (*β − γ*) *t*.

*δ* **estimation**. The values of the hidden asymptomatics and *δ* are determined using a grid search and then choosing the values which provide the best fit. The estimates of *δ* for the 186 districts were 0.34 ± 0.02, which corresponds to a timescale of 4.23 ± 0.48 days. This timescale is comparable to the mean incubation time estimates [9].

*R*_*t*_ **estimation**. The time-dependent reproduction ratio, *R*_*t*_, which is an effective measure of the current status of the spread of the infection and its containment was obtained from the ratio *β*(*t*)*/γ*(*t*).

### Linear trend analysis

The various epidemic parameters we estimated were analysed for the presence of a significant trend. This linear trend analysis was performed using the Matlab module regstats and the significance values were obtained using stats.tstat.pval.

### Travel accessibility of districts

The ease of travel-access to the districts was defined using the *pre-pandemic* airplane and train connectivity. The international and domestic flight traffic to the different airports was gathered from the Airports Authority of India website (*https://www.aai.aero/sites/default/files/traffic-news/Mar2k20Annex2.pdf*), and it was assigned to the respective districts. The train station with the highest through-traffic in each district was noted using the data from the *ClearTrip* travel website (*https://www.cleartrip.com/trains/stations/list*). A combined metric for travel accessibility was developed where the districts were ranked 1 to 8 and further grouped into four categories - *Large metro districts:* International airports with an *annual* traffic of more than 25, 000 flights (1), more than 5,000 flights (2), with any direct international flights (3), *Metro districts:*more than 5000 *annual* domestic flights (4), any domestic flights (5), *Urban districts:* more than 50 trains stopping in the district (6), more than 20 trains (7), and *Sub-urban districts:* all districts with other quaternary means of reaching (8).

## Results

### Hotspots are not contiguous

During the April 15-August 31 of 2021, the number of COVID-19 infections in India rose continuously from around 1, 000 daily new infections to around 80,000 daily new infections. The initial epicenters were in the large metropolitan cities such as New Delhi (from the state of Delhi) and Mumbai (from the state of Maharashtra). With lockdowns and non-pharmaceutical interventions, the case burden in these cities reduced, but there was a concomitant rise of COVID-19 cases in many smaller districts. The active-case burden diffused out of large metro districts. However, mapping the hotspot districts in the geographic map of the India certainly does not immediately reveal how the infections increased or spread. An example of the development of hotspots in the state of Maharashtra is shown in Figure 2. One may immediately notice two exceptions - that newer hotspot districts need not be contiguous with the existing hotspots and further some of the contiguous districts did not develop into hotspots. Thus it is apparent that a plain spatial diffusion model is not sufficient to address the hotspots that appear non-contiguously. We needed an alternative framework to interpret the development of the hotspots.

**Figure 2:**
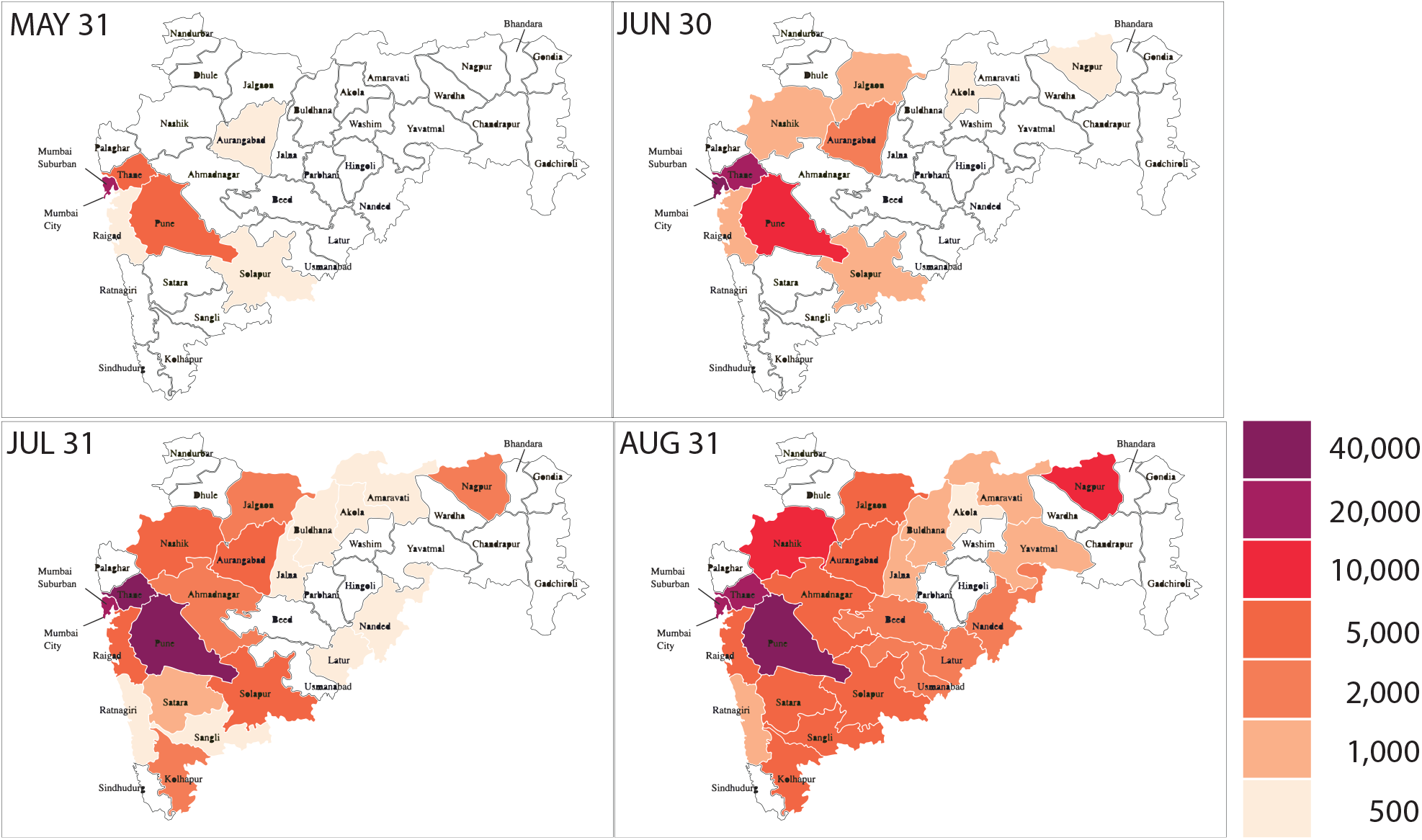
Evolution in time of the pandemic over the districts as seen in the geographic map of Maharashtra. The active case burden is approximated to the nearest cluster indicated in the color-bar. One can see that the the spread of the infections is at times continuous, and on some other occasions begins in districts disconnected from the current hotspots.

### Correlating hotspots with international travel accessibility

Since COVID-19 infections started with import of cases, we asked if the downstream development of newer hotspots is also related to the ease of accessing a district from an international perspective by various means of transport. We ranked the districts 1 to 8 depending upon the level of accessibility, and further categorized the districts as large metro, metro, urban and sub-urban districts using a hierarchical accessibility by international airports, domestic airports, train hubs and other quaternary means of transport, respectively (**Methods**). In Figure 3, we show the number of active COVID-19 cases during the period May 15 - August 31 across the districts in four broad accessibility categories. The figure clarifies that the hotspots in each state, represented by a sector, spread radially outwards suggesting a spread away from the districts with international airports towards districts which are only accessible only via trains. The states where the most accessible regions have not yet recorded a lot of infections, did not also a downstream spread of infections into their remote districts. The figure highlights the significant role of importation of the cases, and a weak or poor-mixing across different states initially due to lockdowns, followed by quarantines upon inter-state travels during phased release of the lockdown.

**Figure 3:**
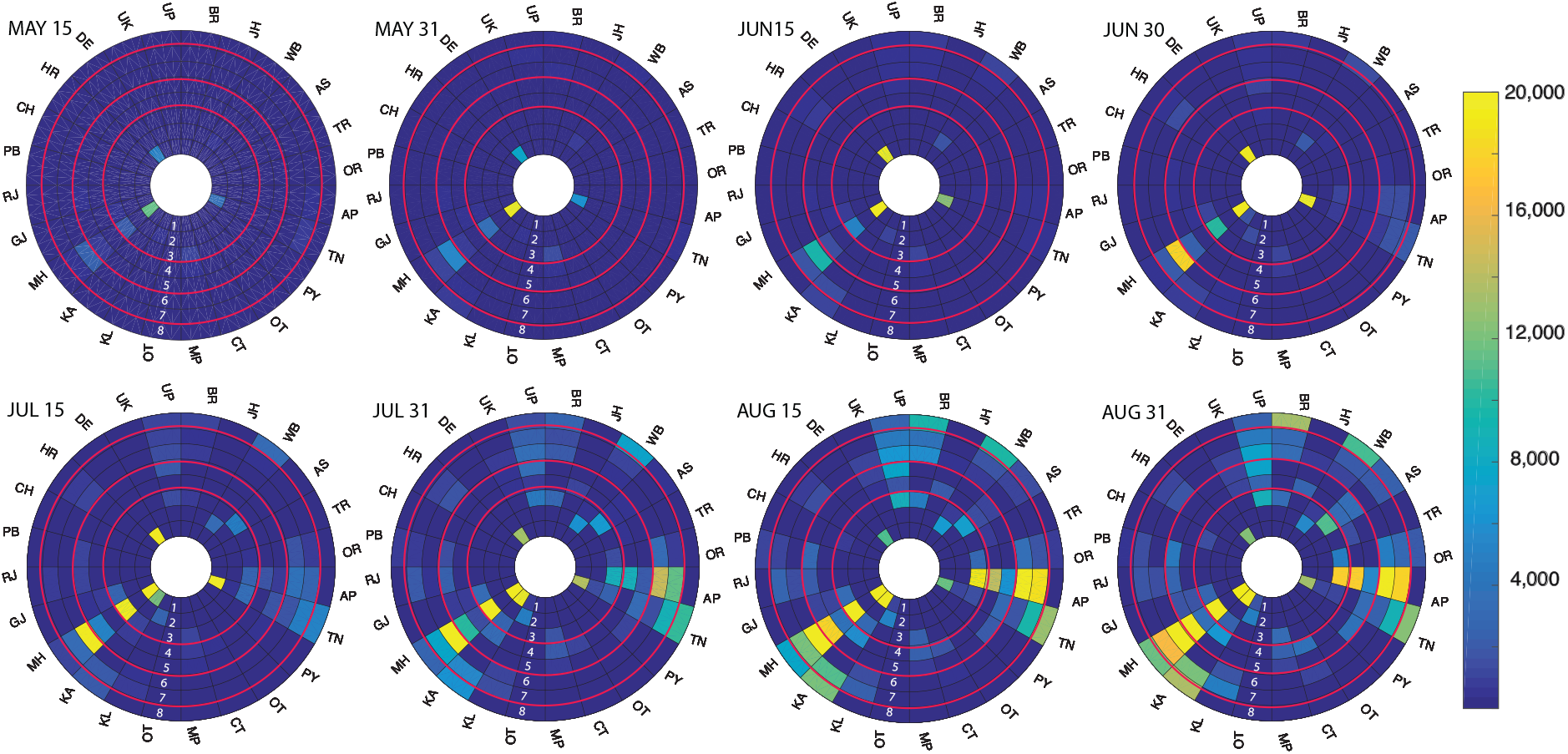
Evolution in time of the pandemic over the districts. Each sector represents the state denoted by its 2 letter code, such as DE for Delhi, MH for Maharashtra. The 8 different layers from inside to outside are the ones in the decreasing order of accessibility. These 8 layers are further clustered as those having international airports, having only domestic airports, connected only by trains and have no train or airplane connections, and are separated by the red-circles. In each state, the hotspots continue to move outwards as the time progresses. Two segments labeled OT are meant other states, which are not included in the analysis.

### The pandemic-burden shifted away from larger metros in the first wave

As the infections continued to grow in numbers as well as spread across the country, it is important to know where the current center of the disease burden is, and if there is a predictable shift in its position. We defined the current center of active case burden by weighing the number of currently active cases with the accessibility level (1 to 8) of the district as (Σ_*districts*_ *I*_*district*_ × accessibility)/(Σ_*districts*_ *I*_*district*_). The result shown in Figure 4 suggests a continuous and smooth drift of this center of active cases away from the large metro districts towards least accessible districts. The districts which are not easily accessible, are also usually the ones with lesser health care resources. The result underscores the need for planning and deploying resources towards regions which do not have adequate testing and critical health care support.

**Figure 4:**
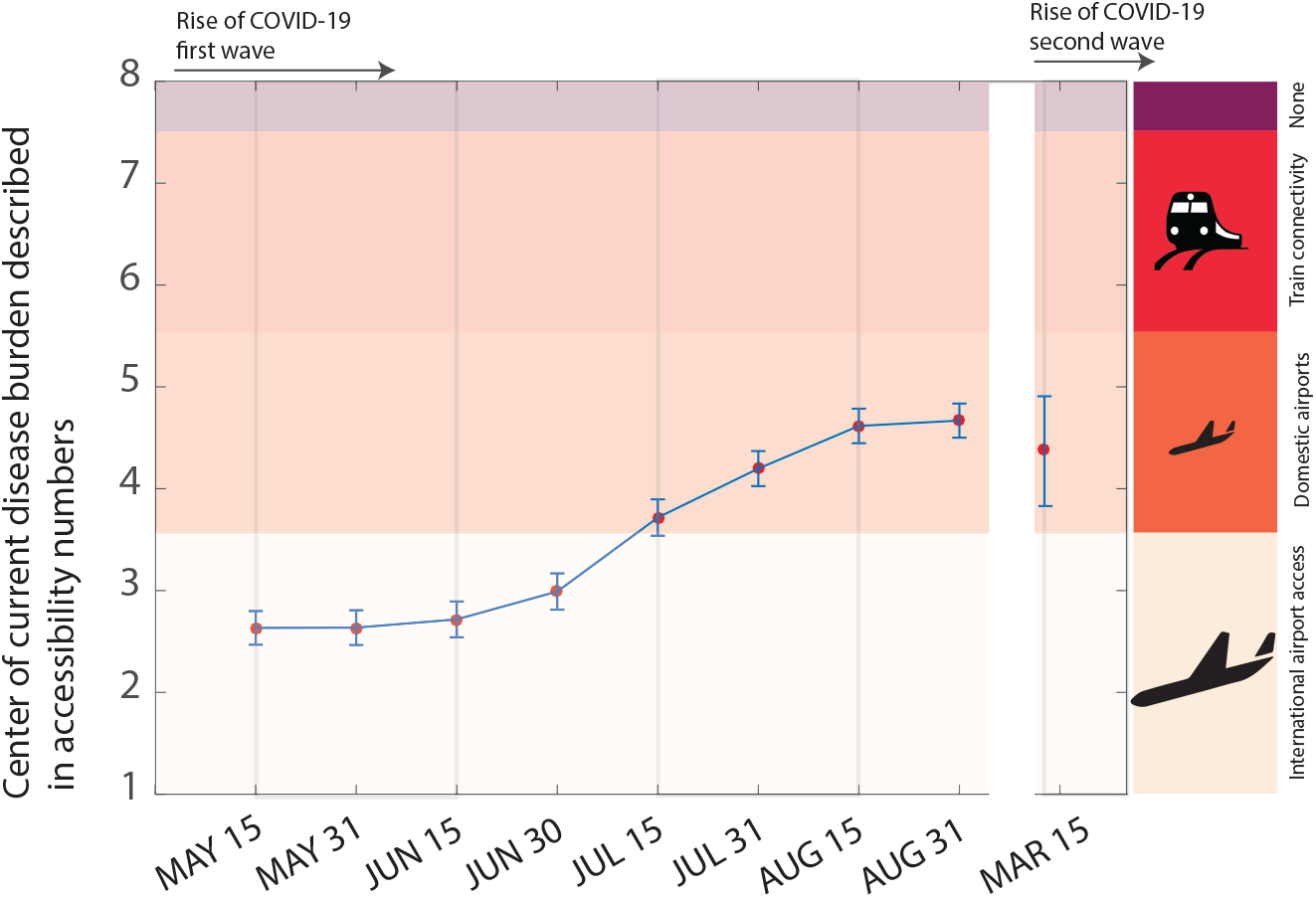
The current burden of the active cases is calculated by weighing the active cases in each district by its accessibility code (1-internationally accessible large metro; 8-least accessible district, as indicated in Figure 3). One can see a clear shift of the case burden away from the large cities. The accessibility of the districts through international, domestic airports and trains are indicated. Also shown is the infection burden as of March 15, 2021 at the beginning of the second wave of COVID-19 infections, which in a way seems to continue with the spread of infections in small and hard to access districts.

The peak of the first wave of infections was reached in mid-September 2020, and the number of infections in the various regions we studied decreased without any further geographical spread. In mid March 2021, the alarm about the second wave of infections began as over 40,000 daily new cases were reported. The epidemiological data from this early stages of the second wave shows an interesting pattern that the districts with the highest number of infections are mainly with medium or low accessibility (Figure 4). As the second wave progresses, these patterns need to be reassessed to understand or validate the role of mobility across the districts.

### Infections continued to rise, but with lower Reproduction numbers

In addition to this geographical spread, we also analyzed the temporal trends in the epidemic parameters from the different regions. During the rising phase of the first wave of infections even as the number of newly reported cases and currently active cases continued to rise (Figure 5A), one can notice from the slopes that the rate of infection rise continues to decrease in districts of all levels of accessibility. The time varying reproduction number (*R*_*t*_) estimates from our analysis over the May 15-August 31 period which witnessed phased release of lockdowns suggest values which are much less than the basic reproduction number estimates between 2.5 to 4. The average reproduction number across the districts of different accessibility showed a decreasing trend (Figure 5B). A linear trend analysis suggested the large metro and sub-urban districts have a reduction in *R*_*t*_ with p-Values of 0.0001053 and 0.0067 respectively. While the average reproduction numbers appear to reduce even in metro and urban districts, the trends were not significant (p-Values of 0.4634 and 0.4618 respectively).

**Figure 5:**
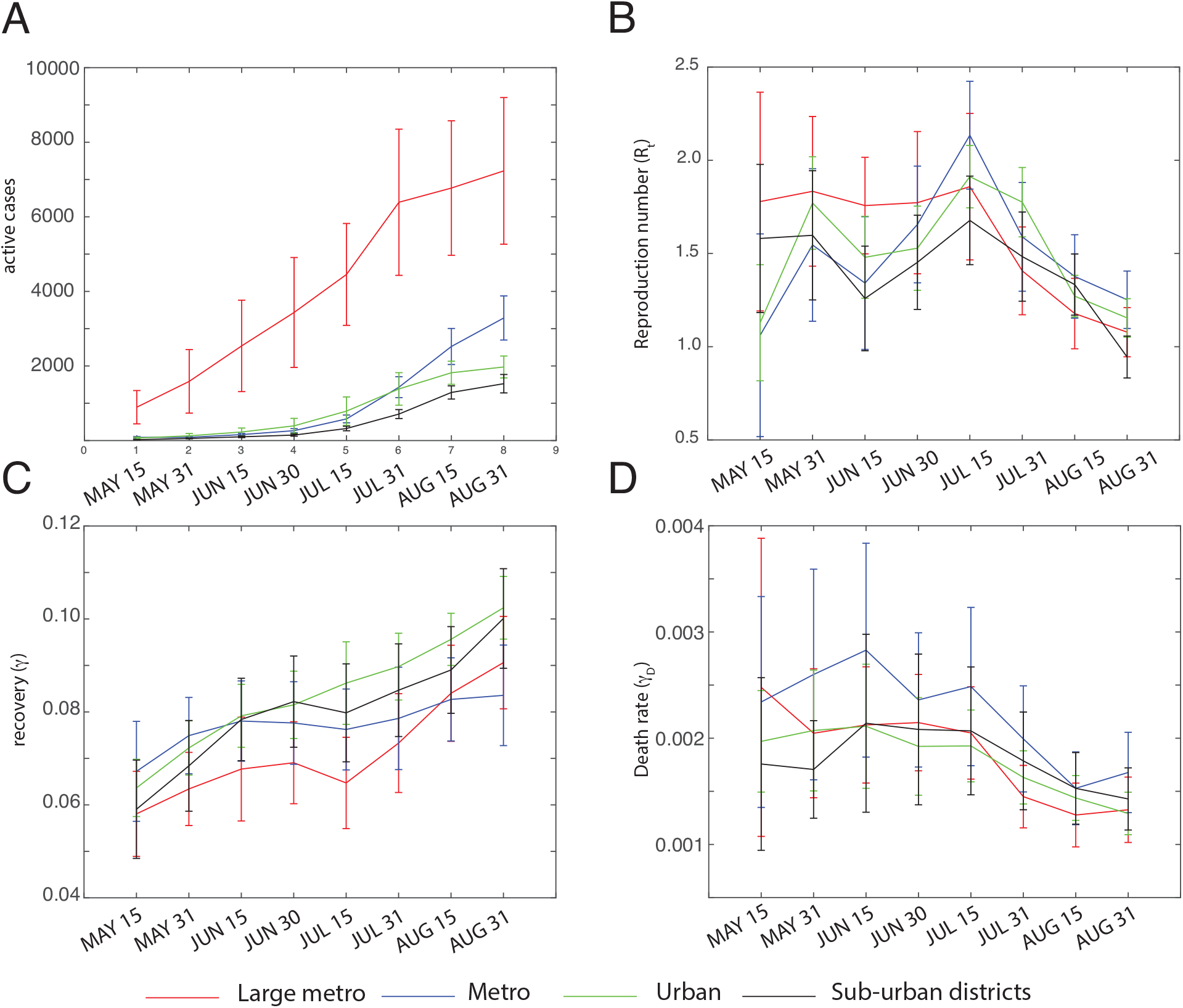
The trends in different epidemiological parameters *A*. Number of daily new infections, *B*. Reproduction number, *C*. Recovery rate (*γ*) *D*. Death-rate (*γ*_*D*_) are shown for the districts categorized by their accessibility is shown. The trends across the different districts groups are consistent. The reproduction rates, recovery and death rates are all improving, despite the increase in the new infections.

In order to distinguish if the slow down of infections occurred because of a herd immunity or because of the changes in behavioural patterns, we examined the infected percentage (=cumulative infections/population × 100%) of the population at which the *R*_*t*_ dropped below one. The infected percentages vary significantly, for example in the two contiguous districts of Mumbai and Pune, these values were 0.25% symptomatic and 1.08% including asymptomatics in Mumbai and 0.7% symptomatic and 2.21% including asymptomatics in Pune, which suggests that the slowdown of infections is achieved because of changes in behavioural patterns and an exercise of caution was still required to maintain the pandemic in control.

The trends in the reproduction numbers in the second wave of the COVID-19 infections need to be examined when the sufficient number of infections are built over time to yield reliable predictions of the epidemic parameters across the different districts. Further, any such analyses will also need to account for the presence role of different mutants.

### Recovery and death rates improved

The time to recovery from the time of appearance of symptoms for the symptomatic patients (1*/γ*) estimated for the 186 districts as 16.18±1.01 on May 15 was consistent with the 24.7 (22.9-28.1) that was reported in the early days of infections from China.[10]. However, by August 31, the recovery time showed an improvement as (*γ*) decreased to 10.08 0.69 on August The time variation of recovery rates (*γ*) is shown in Figure 5C, and the trend analysis suggested an increase in the recovery rates with p-values 0.2667×10^−12^, 0.5504×10^−3^, 0.9180× 10^−37^, 0.1250×10^−16^ in the large metro, metro, urban and sub-urban districts. Similarly death rates *γ*_*D*_ which is a product of the case fatality rate (reported as *≈* 2% in India, and consistent with an *≈* 10 days time to death) with the rate of death also showed an improvement over the time with p-Values of 0.0010, 0.0122, 0.6272 × 10^−3^, 0.2789 respectively in the large metro, metro, urban and sub-urban districts (Figure 5D).

### Comparison with other studies

In an analysis of the spread of SARS across countries,[11] it was realized that a smooth spatial diffusion of pandemic which was sufficient to describe Black Death[12] could not address a quicker reach of the epidemic to many cities. It was demonstrated that the time taken for the epidemic arrival time to the different cities was correlated not to the geographic distance from the epicenter, but rather to an effective distance defined by the degree of connectivity between the different international airports. The work thus introduced the concepts of development of epidemics that go beyond simple spatial diffusion.[12] In a cross-sectional analysis, the development of case-load of the infections in the different hotspots in China was correlated to the outflow of traffic from Wuhan in the period January 1 to January 24.[5] In another recent study on COVID-19 in Brazil, the connectivity between different cities via highways was attributed to be one of the reasons for super-spreading across major cities.[13] However, the work did not establish any patterns for the spread of the active-case burden or its flattening,[14] which is extremely important in an epidemic or a pandemic from the public health point-of-view.

While the present work is in line with all three works noted above in term of travel connectivity, there are several unique aspects we discovered in our study of the COVID-19 data from India. The global spread of pandemic from Wuhan to different major international destinations may have followed the similar pattern of effective-distance.[11] However, the emphasis, from the perspective of incidence within a country and critical resource planning is on the granular data within the country. Further, unlike the travel outflow which was obtained by tracking mobiles,[5] the accessibility we use via airplane and train-connectivity data from *pre*-pandemic period is believed to be a surrogate for the degree of connectivity and mixing among the populations. In the *network science* parlance, this is an emphasis on the centrality-measures which define the relative importance of the different cities that are being connected, by-passing the actual connectivity data which is harder to obtain, especially because our study was not limited to a single phase such as the January 1 to January 24.[5] Finally, by developing the hierarchical accessibility network, we could show trends such as the radial spreading out towards remote areas in each state and a smooth variation of the current-case-burden in Figure 4. Figures 3 and 4, in our opinion go beyond the immediate requirements of how the infections are rising in a specific region and serve to provide a wholistic perspective on the development of the pandemic as well to help planning for allocating or re-organizing critical care or test resources towards less catered cities.

## Conclusions

In summary, we show predictable trends of improvement of disease parameters, and spread of COVID-19 infection burden away from easily accessible metros with large international airports. The continuing number of cases, and the predictable drift suggests the need for planning or even reorganizing testing and critical health care resources to districts which are down-stream in international access. A lack of correlation between the slow-down of infections and the extent of current infections suggests that the caution and change in behavioral patterns need to continue if COVID-19 has to be kept in check.

## Data Availability

The work is based on publicly available data. The scripts and data will be made accessible upon request.

https://github.com/meherpr/COVIDIndianDistricts

## Declarations

## Acknowledgements

We acknowledge discussions with Prof. P. Sunthar. MKP and SA acknowledge Aman Sharma and Fauzia Javed for help in gathering the airport and train traffic data.

## Availability of data and materials

The analysis was based on publicly available district-wise infection and death rate available from: https://www.covid19india.org/. The district-wise COVID-19 infection, recovery rates we estimated, the flight and train traffic through these districts, and the Matlab script used for analysis are all available at: https://github.com/meherpr/COVIDIndianDistricts.

## Authors’ contributions

SAnsumali and MKP conceived and designed the study; SAgrawal performed the SAIRD model analysis; HJS collected district accessibility data; SAn-sumali, AK and MKP analyzed the results; SAnsumali and MKP wrote the manuscript.

## Ethics approval and consent to participate

The work is an analysis of publicly available epidemiological data. So no ethics approval is not applicable.

## Consent for publication

Not applicable.

